# When are predictions useful? A new method for evaluating epidemic forecasts

**DOI:** 10.1101/2023.06.29.23292042

**Authors:** Maximilian Marshall, Felix Parker, Lauren M. Gardner

## Abstract

**Background:** COVID-19 will not be the last pandemic of the 21st century. To better prepare for the next one, it is essential that we make honest appraisals of the utility of different responses to COVID. In this paper we focus specifically on epidemiologic forecasting. Characterizing forecast efficacy over the history of the pandemic is challenging, especially given its significant spatial, temporal, and contextual variability. In this light, we introduce the Weighted Contextual Interval Score (WCIS), a new method for retrospective interval forecast evaluation. The WCIS reflects the potential utility of predictions, resulting in a score that is easily comparable across different pandemic scenarios despite remaining intuitively representative of the in-situ quality of individual forecasts.

**Methods:** The central tenet of the WCIS is a direct incorporation of contextual utility into the evaluation. This necessitates a specific characterization of forecast efficacy depending on the use case for predictions, accomplished via defining a utility threshold parameter. In essence, changes in forecast accuracy beyond this threshold do not map to changes in the utility of a prediction. This idea is generalized to probabilistic interval-form forecasts, which are the preferred prediction format for epidemiological modeling, as an adaptation of the existing Weighed Interval Score (WIS).

**Results:** We apply the WCIS to two different forecasting scenarios. The first assesses the performance of facility-level COVID-19 hospital bed occupancy predictions for the state of Maryland during the Omicron wave, and the second evaluates state-level hospitalization forecasts drawn from the COVID-19 Forecast Hub. We use these applications to demonstrate the parameterization of contextual utility, compare the WCIS to the WIS, and explore the utility of the WCIS.

**Conclusions:** The WCIS provides a pragmatic utility-based characterization of probabilistic predictions. This method is expressly intended to enable practitioners and policymakers who may not have expertise in forecasting but are nevertheless essential partners in epidemic response to use and provide insightful analysis of predictions. We note that the WCIS is intended specifically for retrospective forecast evaluation and should not be used as a minimized penalty in a competitive context as it lacks statistical propriety.

## 1 Background

### 1.1 Introduction

Given the devastating impact of COVID-19, and in the face of future pandemic threats, it is incumbent upon the epidemic forecasting community to deploy prediction tools that provide meaningful and actionable utility to those who need them. An important piece of this effort is candid retrospective evaluation of the utility of forecasting during the COVID-19 pandemic. In this light, we present a new probabilistic forecast evaluation method, the Weighted Contextual Interval Score (WCIS). It is a relative metric that encodes a simple question. How useful could a forecast have been where and when it was made? Unlike other scores, the WCIS is designed specifically as a retrospective way to judge whether or not forecasting could have been useful. It is not intended for real-time model ranking and ensemble construction. Instead, the WCIS is meant for broader pandemic preparedness efforts, for taking an honest look at how helpful forecasts could have been and thus potentially could be in the future. Despite the high spatial and temporal variability of pandemic scenarios, the WCIS evaluates forecasts in a comparable and communicable way by scoring them as a function of their potential utility.

The advent of the COVID-19 pandemic precipitated a massive public health response, including a significant modeling effort [1, 2]. In the United States, this quickly resulted in the formation of the COVID-19 Forecast Hub, a repository for short-term pandemic predictions [3]. Similar to prior collective forecasting efforts focused on seasonal influenza, dengue, and Ebola, the Forecast Hub solicited predictions from a large and diverse group of modelers, synthesizing their submissions into ensemble forecasts of COVID-19 cases, deaths, and hospitalizations. These outputs were provided to the United States Centers for Disease Control and Prevention (CDC) for policy making and dissemination to the public [4–9]. In addition to modeling efforts like the Hub at the regional level, COVID prompted a considerable amount of more granular forecasting, such as predictions for individual medical facilities [10, 11]. Despite this abundance of pandemic modeling, translating short-term epidemiological forecasts into applicable, actionable, and insightful decision-making remains a significant challenge [7, 12–17]. Understanding whether or not a forecast could have been useful requires understanding the conditions in which the forecast was made. It also requires knowledge about the type of decision the forecast would be used to inform. The WCIS was designed around these two requirements. It uses a utility-based normalization scheme to enable intuitive and meaningful comparison of forecast quality despite potentially dissimilar prediction contexts.

### 1.2 Motivation

Probabilistic predictions are preferred in many disciplines, including the epidemic forecasting community. Unlike single outcome “point” predictions, probabilistic forecasts convey the uncertainty of the underlying model. This is particularly important given the difficulty of correctly interpreting a quickly-evolving pandemic [7, 18]. The extant Weighted Interval Score (WIS), an error metric for quantile forecasts that approximates the Continuous Ranked Probability Score, is the primary method used to evaluate Forecast Hub submissions [19, 20]. As summarized by Bracher et al., “the (Weighted Interval) score can be interpreted heuristically as a measure of distance between the predictive distribution and the true observation, where the units are those of the absolute error” [19]. The WIS is an effective metric for real-time prediction scoring, model comparison, and ensemble forecast creation [20]. However, the WIS is limited in its ability to be used for intuitive forecast utility analysis, in particular because the score is scaled on the order of the prediction data [19]. Retrospective pandemic evaluation involves comparing scenarios of highly different scales. One example of such a comparison would be between regions with large baseline differences in data magnitudes, such as highly vs sparsely populated regions (as in the Forecast Hub). Another situation where scale-related contextualization is essential to consider is the comparison of periods of high vs low epidemic activity (surge vs non-surge). In fact, both of these spatial and temporal scaling challenges are often necessary to consider at the same time (see Additional file 1: Section 1.1 for motivating examples of these issues drawn from state-level pandemic scenarios in the United States).

The WCIS is an adaptation of the WIS that is framed around the two following ideas. First, any meaningful measurement of forecast quality must arise from the context into which predictions are disseminated. In other words, a useful forecast improves real-time knowledge and/or decision-making capabilities. The reverse also holds: a forecast is not useful if it is incapable of (or if it provides information detrimental to) gaining real-time information or improving decision-making. Second, for the purposes of enabling the comparison of forecast performances in disparate scenarios without post-processing, a helpful score should be a relative metric. Taken together, these two concepts informed the central idea of the WCIS: that a consistently meaningful score must have endogenous contextualization. In essence, the WCIS normalizes forecast performance as a function of the ability of the forecast to be used in the specific environment in which it was made. This way, despite potentially occurring in radically different spatial and temporal scenarios, individual evaluations can be meaningfully compared to others.

Before moving to its technical basis and formulation in the next sections, it is necessary to address the intended purpose of the WCIS, and similarly, tasks for which it should not be used. The WCIS is not a statistically proper score (see Additional file 1: Section 1.3), which means it should not be used in competitive forecasting contexts. In these situations, such as real-time evaluation of COVID-19 Forecast Hub submissions, scores that are not statistically proper have the potential to be gamed [21]. Again, the WCIS is not designed for and should not be used for such purposes. It was not created to replace the WIS, which functions well for real-time forecast scoring and ensemble generation. Instead, the WCIS is designed to reflect relative forecast quality using a flexible and contextually specific parameterization of utility. As is demonstrated in the test cases below, this results in a score that is not just intuitively interpreted, but is easy to compare and convey visually. We believe that these attributes are highly important in the context of pandemic preparedness efforts, given the need to more strongly connect the modeling and policy-making spheres of the public health community. Decision-makers need to be able to assess whether or not forecasting has the capacity to positively contribute to pandemic response. We believe that the WCIS enables an intuitive and flexible exploration of this question.

### 1.3 Review of the Weighted Interval Score

The Weighted Contextual Interval Score (WCIS) builds directly from the Weighted Interval Score (WIS). Bracher et al. [19] provide an excellent explanation of the mechanics of the score and its applications in epidemiology, and we endeavor to use the same symbology whenever possible. For brevity, the entire WIS formulation is not reviewed here, but the key elements (that are also important pieces the WCIS) are necessarily summarized:

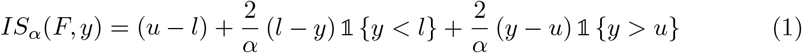

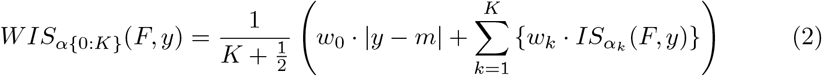

- We assume a submission of *K* interval forecasts drawn from a predicted distribution *F*, a probabilistic representation of the target variable. Each of the *K* forecasts represents a (1 − *α*_*k*_) prediction interval (PI). These intervals are delineated by their lower and upper bounds *l* and *u*, the 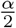 and 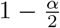 quantiles of the predicted distribution, respectively. For example, a 95% interval would be represented by an *α*_*k*_ of 0.05, its lower and upper bounds defined by the 0.025 and 0.975 quantiles of *F*.
- A predictive median *m* (point prediction) is submitted, and the true target value *y* is known.
- For each interval *k* ∈ {1, 2, …, *K*}, an individual Interval Score (IS) is calculated, penalizing both the width/sharpness of the interval: *u* − *l*, and (if necessary) the amount by which the interval missed the true value: 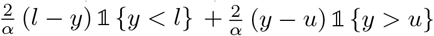 [21]. Note that the “miss” component is scaled by the inverse of *α*, thus narrower prediction intervals are penalized less for missing than are higher confidence submissions.
- The WIS is a weighted average of each of the *K* Interval Scores and the absolute error of the predictive median, with the weights *w*_*k*_ used for the average corresponding to 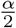 for each interval.

## 2 Methods

### 2.1 Contextualizing Point Forecasts

Although the WCIS (like the WIS) is an interval score, it is framed around a point score that we call the Contextual Relative Error (CRE). The CRE maps the absolute error of a point forecast *x* to its contextual utility. This is achieved by specifying *δ*, the utility threshold parameter. (Note that *δ* is the only parameter in the WCIS formulation that does not already appear in the WIS score).

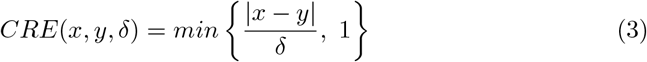

*δ* is the magnitude of the absolute error above which a forecast loses its utility. The CRE is so named because instead of mapping to the distance between a predicted value and its target like absolute error, it maps to an interval from 0 to 1. A score of 0 indicates a forecast with maximum possible utility (with an absolute error of 0), and a score of 1 indicates a useless forecast (with an absolute error of *δ* or more). See panel *(a)* of Additional file 1: Fig. S1 for a graphical representation of the CRE. An important feature to note is the “plateau” of the metric when the absolute error exceeds *δ*. This might seem problematic, given that beyond the *δ* threshold the absolute error is capable of increasing without any commensurate increase in the CRE. This is, in fact, the desired behavior of the CRE and warrants a slight re-framing of perspective. Selecting *δ* requires, when applying the CRE (and the WCIS, as it is a generalization of the CRE from point to interval scores), identification of a practical limit for how a forecast is used or interpreted in a particular context or for a particular purpose. For example, in many scenarios we have a finite capacity to respond to an expected outcome. If the “demand” imparted by an incorrect forecast exceeds that capacity, we are unable to alter our response despite an apparent increase in need. Therefore, an incorrect forecast with an absolute error of 2*δ* wastes exactly as many resources as a incorrect forecast of magnitude *δ*, where *δ* precipitates the maximum allocation in response to the forecast. A different way to interpret *δ* is as an “absorbable error magnitude.” The test cases later in the paper frame *δ* this way, wherea decision maker has limited capacity to recover from plans made according to forecasted outcomes. If the forecast is wrong enough that it precipitates an action that cannot be recovered from, such a forecast has met or exceeded the *δ* threshold.

Note that *δ* is both a normalizer and a limit. Thus a forecast with an absolute error greater than *δ* is not at all useful, and a forecast with an absolute error less than *δ* is evaluated as a ratio of *δ*. This gives the CRE (and the WCIS) the ability to provide information about both forecast quality and how frequently forecasts are useful, which, as demonstrated later, is helpful for intuitive analysis and performance visualization.

### 2.2 Contextualizing Interval-Form Forecasts

We begin by introducing the Contextual Interval Score (CIS). The CIS is both a probabilistic extension of the Contextual Relative Error, and a contextualized version of the Interval Score. Like the CRE, it maps a forecast’s error to the *δ*-parameterized utility space, and like the IS, it generates a score for a single interval forecast. (In fact, the CIS can be equivalently formulated in two different ways, based on either the IS or the CRE. For brevity, we use the IS-based formulation here but, particularly if more intuition about the score is desired, we suggest referencing Section 1.2 in Additional file 1 for the explanation of the CRE-based formulation.)

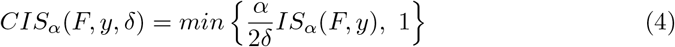

The WCIS is the simple average of the CIS across all *α*-intervals and the CRE of the predictive median *m*:

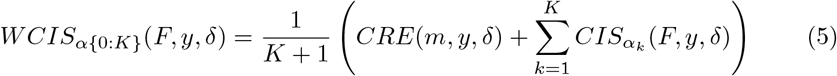

Note that we still retain the descriptor “Weighted” in the WCIS title even though there are no weights directly included in its formulation, whereas each component of the WIS is multiplied by 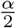. However, in our formulation, the same weights are effectively applied directly to the individual constituent CIS scores. Instead of the “miss” components of the score being multiplied by 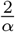, the “width” term is scaled by 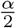. Thus when the average is taken to create the WCIS the scaling effect is the same as the WIS, but the weights are applied in this way because it preserves the interpretability of the individual single-interval CIS components as described above. Another notable difference is the WCIS uses *K* + 1 for the denominator of the average (unlike 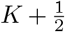 in the WIS) because like the single-interval components, the predictive median component of the score has a maximum penalty of 1. This, and the bound on each CIS term, means the WCIS also takes values only on the interval from 0 to 1. Note the natural equivalence between the WCIS for interval forecasts and the CRE for point forecasts, which mirrors that between the WIS and the absolute error. In both cases, the interval scoring method preserves the behavior and intuitive interpretation of the corresponding point forecast technique.

## 3 Results

The WCIS is expressly intended to be a flexible scoring method and as such there are many possible and highly variable ways to apply it. We use this Results section to present two demonstrative use cases. Both scenarios evaluate COVID-19 hospitalization forecasts, but each works at a different scale and uses a necessarily different *δ* formulation. The first scenario applies the WCIS to results from a multi-facility-level forecasting model. We use this first application primarily to develop the intuition for the *δ* selection process. We show via a direct demonstration how *δ* can be chosen to represent contextually specific utility as a function of time-varying data, and explore how the choices made during this parameterization map onto the output of the WCIS.

Since this section focuses more on the WCIS formulation and less on interpreting the real-world applicability of the predictions, we use forecasts from a model developed in-house. Conversely, the second test case evaluates four weeks ahead predictions from the COVID-19 Forecast Hub’s ensemble model, examining hospitalization forecasts from May 2021 to May 2022 [3]. This period includes both the Delta and Omicron variant waves and allows for a larger exploration of the utility and communicability of the WCIS. Data for these analyses are sourced from the COVID-19 Reported Patient Impact and Hospital Capacity by Facility dataset for the first section and from the Forecast Hub’s repository for the second [22, 23].

### 3.1 Facility-level Analysis (First Test Case)

As introduced above, our first test case evaluates a facility-level hospitalization model. More specifically, the model forecasts daily COVID-19 bed occupancy, for each individual hospital in Maryland, from one to twenty-one days out, from July 2021 to July 2022. Because our *δ* selection reflects capacity management within the three-week forecast window, we only use hospitals listed as “short-term” type (this excludes longterm and pediatric facilities) and for relevance only include facilities that had at least ten COVID-19 patients at some point during the time range specified. The particular time range used was chosen because contextualization is vital when comparing and contrasting scenarios with highly different levels of pandemic activity, and July 2021 to July 2022 includes the Omicron wave in Maryland. This scenario and facility selection yields 42 hospitals with an overall capacity range of 30 beds at the smallest facility to 919 beds at the largest facility.

The model used is a Time Series Dense Encoder, using the prior ninety days for each hospital at each time point to predict the following twenty-one days [24]. For a complete model formulation see Additional file 1: Section 2.1 but in brief, this model type was selected because it is a state-of-the-art general-purpose time series forecaster that is efficient to train and flexible across different covariates, prediction horizons, output types, and loss functions. We note that the purpose of this test case is to explore and explain the formulation and application of the WCIS. Thus, we developed this relatively basic model in order to apply the WCIS to a facility-level scenario, not to refine a specific method for forecasting hospitalizations. The predictions from this section are not necessarily indicative of those performed in real time. Because the data used for training and scoring this model may contain retrospective corrections of errors that were present in the real time data, it has the potential for higher performance when compared to an equivalent in-situ forecaster.

The *δ*-parameterization used for this analysis is intended to characterize the capacity of each facility to absorb an incorrect allocation of COVID-19 bed space based on a flawed forecast. We assume that capacity allocations are made at forecast time, under the in-situ assumption that forecasts perfectly reflect future outcomes. Thus, the *δ* value represents an achievable capacity correction during the time interval separating the making of the forecast and the realization of its true target value. For example, the *δ* value for a seven-day-ahead forecast for each facility is the amount of COVID-19 beds that each individual hospital can add or take away over a week. Specifically, this *δ* is determined as follows. The daily capacity change for each hospital is calculated as the mean of all single day, non-zero capacity changes over the entire available time series for each facility. *δ* for a particular forecast is then set as the product of the forecast horizon and the facility-specific daily change capacity. This means that the further out a forecast is, the larger (and thus more forgiving) the delta value is, based on the idea that the more time a facility has to respond to a poor allocation of resources, the greater the magnitude of the response can be. Please note that the particular formulation chosen here is not intended to provide an assessment of forecast quality outside the utility scenario posited by the assumptions given above. However, it demonstrates an important capability of utility threshold selection: *δ* can be a defined as a dynamic function of data that can change in time and space. Since contextually meaningful forecast utility varies significantly over these same dimensions, a broadly applicable and interpretable score must be similarly adaptable.

Using Figures 1 and 2, we are able to interpret some important aspects of how this selection of *δ* maps onto the scoring of our facility-level model. First, consider the relationship between the breadth of the confidence intervals and the *δ* region in Figure 1, which visualizes a single facility. The larger prediction intervals for the fourteen day-ahead forecasts indicate less model certainty than those of the two day-ahead predictions and, all else equal, would yield a worse score. However, *δ* is significantly higher for the fourteen day-ahead scenario, given the assumption that facilities have more time to adapt to inaccurate forecasts over longer horizons. This results in generally better performance for the fourteen-day model. However, there remain in the fourteen-day scenario several forecasts that still receive a high penalty despite the more forgiving *δ* parameterization. Note that these instances tend to occur when the forecast median approaches or exceeds the utility threshold. Moving to Figure 2, we can see that these trends are also visible in the aggregate performance across all 42 facilities. Comparing the WIS to the WCIS over these instances reveals a relatively linear relationship in the more forgiving scenarios, i.e. non-wave with a larger delta. During the wave, when absolute performance was broadly worse (as evidenced by the WIS values), the *δ*-limit was reached significantly more often. We also draw attention to the differences in marginal distributions that are visible in the scatter plot column of Figure 2. The scaling and limiting action of the WCIS distributes performances significantly more evenly than the WIS (see Additional file 1: Section 2.2 for plots with these marginal distributions included).

**Fig. 1.**
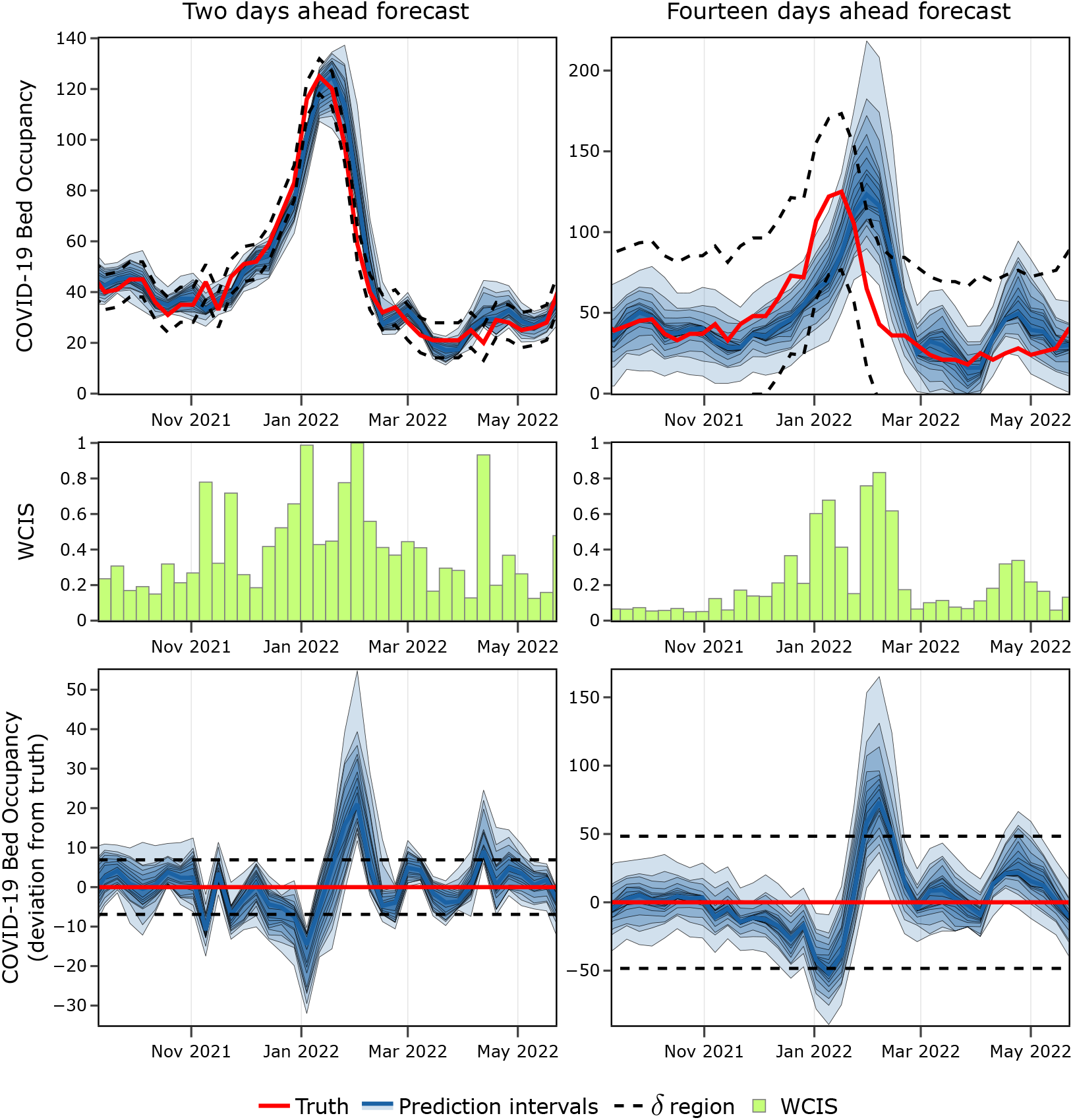
Illustrated here are facility-level forecasts over two prediction horizons for one hospital: the University of Maryland Medical Center. The top and bottom rows both show the same forecasts, truth data, and *δ* (utility threshold) region. The top row displays these values normally, whereas the bottom row shows how far each value deviates from the truth. The middle row displays the WCIS, aligned with the data in the other rows. (Note that the facility-level analysis includes more prediction intervals and more dates than are shown in this figure, the extent of both displayed here are reduced for clarity.)

**Fig. 2.**
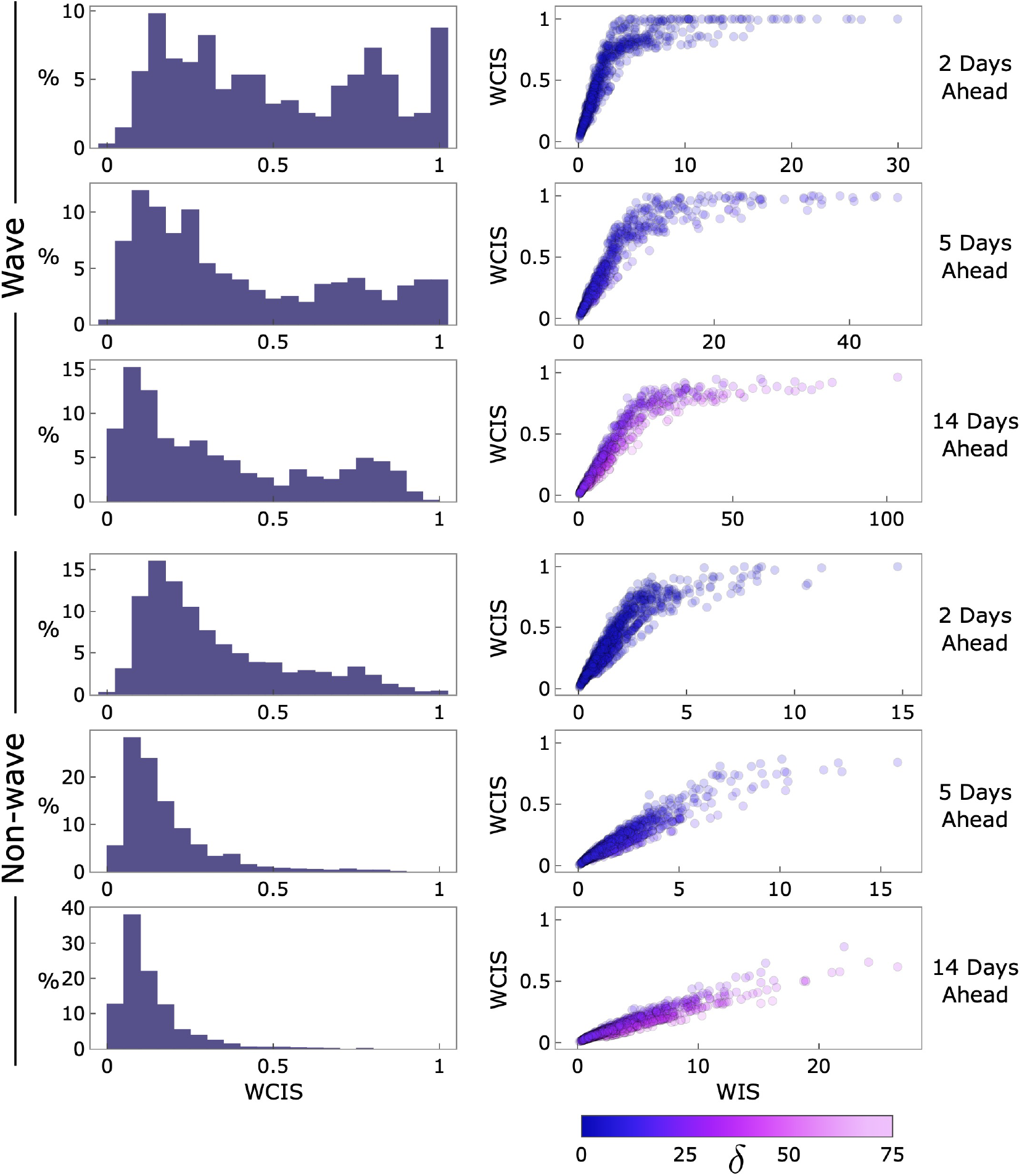
Results in this figure are generated from all 42 hospitals, for all prediction dates in the facility-level model. The top three rows are from forecasts during the Omicron wave, and the bottom three from before and after the wave. We define the wave as lasting from November 14 2021 through May 15 2022, as illustrated in Additional file 1: Fig. S4.

In general, we are able to observe that given a contextually relevant *δ* choice, the score is able to simultaneously convey an intuitive sense of both relative quality and the overall frequency of useful forecasts, as shown in the histograms of Figure 2.

### 3.2 State-level Analysis (Second Test Case)

For this test case, we apply the WCIS to real-world predictions drawn from the Forecast Hub, asking how much contextual utility hospitalization forecasts provided at the state level from May 2021 to May 2022 [3]. (Note that Forecast Hub hospitalization predictions were performed at daily resolution, but for the sake of visualizing a longer-term analysis we aggregate to and evaluate at weekly totals.)

The WCIS always requires a specific interpretation of the use-case for forecasts in the selection of the utility threshold *δ*. Similar to Section 3.1, we choose to assess hospitalization predictions as a function of potential capacity changes. However, we assume a different decision-making scenario for hospital capacity at the state level than for its facility level counterpart. Due to the disaggregate decision-making apparatus across statewide hospitals and the inherent institutional inertia that must be overcome for larger scale change, we take a more conservative approach to estimating the absorbable error magnitude. Specifically, *δ* is the 0.9 quantile of the prior deviations in each state’s hospital bed capacity over the prediction horizon of the forecast. We assume prior bed capacity deviations are indicative of a state’s capacity to make changes, and that it is more difficult to make changes over a shorter timeline. Thus, any deviation over a shorter-term horizon can also occur for longer term horizons, but not the reverse. For example, when examining one week ahead predictions, only historical capacity changes over the course of a single week are considered. For four weeks ahead predictions, capacity changes for one, two, three, and four weeks ahead are considered. Finally, the 0.9 quantile is selected as the threshold under the assumption that states are not necessarily able to repeat their largest historical deviations, but can approach them. To be clear, this choice of *δ* is a heuristic for the amount of resource allocation, staffing changes, and other matters that hospitals might practically accomplish in response to an assumed change in pandemic dynamics. It is intended to demonstrate the WCIS given a reasonable, data-driven parameterization of forecast utility. Namely, a response predicated on a forecast outside the *δ*-range as defined here would require corrective action of a magnitude that could not be reasonably expected over such a forecast’s prediction horizon.

WCIS performance results for four weeks ahead state-level hospitalization predictions are demonstrated in Figure 3. Since the WCIS was designed primarily as a way to meaningfully evaluate and compare forecasts in disparate contexts, we can easily use it to observe several important aspects of hospitalization forecasting performance. For example, during surges and declines, forecast utility decreases substantially. We can intuit that this is a consistent trend across different locations both by directly observing the large central grid and by examining the lower, spatially averaged array of the figure. In contrast, if we examine the right-side, temporally averaged array, we observe that there is less variability in space than there is in time. Thus, by making an up-front determination about what constitutes a useful prediction (performing the *δ*-parameterization), we are capable of making, displaying, and intuitively evaluating forecasts. This allows, given a well-informed choice of *δ*, for meaningful overall analysis without needing to repeatedly delve into the specific circumstances during which each forecast was made. Without contextual normalization, conveying informative and comparable performance would be much more challenging. This capability, demonstrated by the ease of interpreting Figure 3, is the overall aim for our creation of the WCIS. It permits substantive and easily interpretable performance evaluation.

**Fig. 3.**
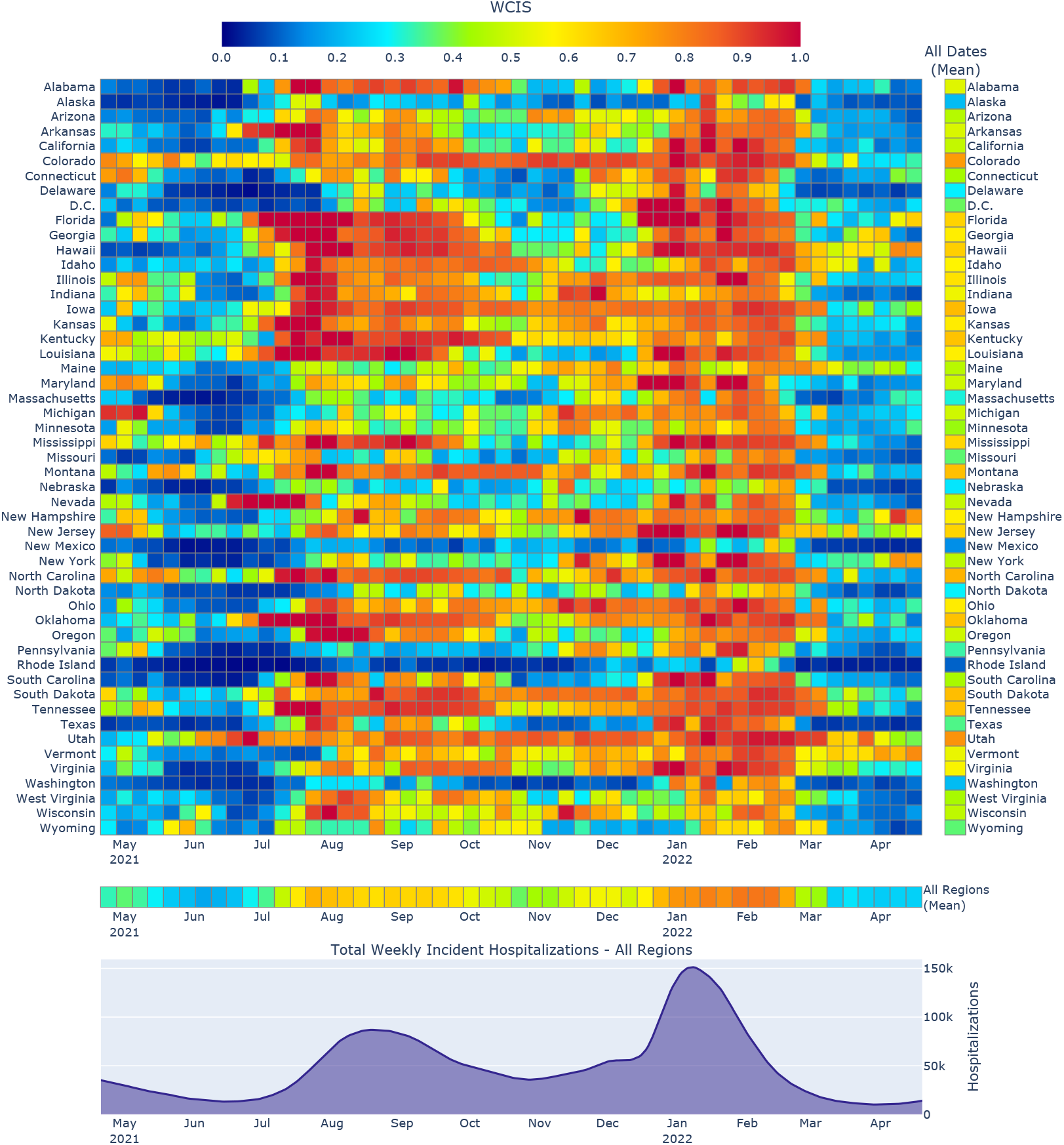
Heatmap of the WCIS for 4 week ahead hospitalization forecasts, performed by the Forecast Hub’s ensemble model. The central and largest grid shows the most granular results: region- and time-specific performance. On the right and lower sides of the grid are average performances over time and space, respectively. The shaded line plot at the bottom of the figure is the target hospitalization variable aggregated across all regions. Note that its domain is aligned exactly with those of the time-dependent heatmaps above, to provide insight into the trends of the overall pandemic alongside the more granular information in the heatmaps. (See Additional file 1: Section 3 for heatmaps over differing prediction horizons).

## 4 Discussion

The WCIS is framed around our belief that a useful forecast contributes meaningful and/or actionable information given uncertain future outcomes. Determining whether or not forecasts accomplish this necessitates an explicit definition of utility. This brings up an important philosophical difference between the WCIS and other techniques. The WCIS formulation, centered around a user-defined utility threshold *δ*, arises from our assertion that there will never be a one-size-fits-all solution for assessing and comparing short-term forecast quality. One must always consider prediction context and purpose lest standard metrics tell a misleading story. Additionally, different forecast use-cases yield different judgments of predictions. The helpfulness of a model that predicts rainfall, for example, will be judged very differently by a user deciding whether or not to bring an umbrella on a walk as compared to a user deciding whether or not to issue regional flood warnings. An incorrect forecast of light rain with a realization of heavy rain is good enough for the first user but may be catastrophic for the second. Again, forecast purpose is essential to consider. The WCIS ensures this by building a definition of forecast utility directly into the formulation of the score.

The core of the WCIS is the combined normalization and thresholding imposed by the *δ* parameterization, which incorporates a vital aspect of real-world forecast utility. Namely, past a certain point, changes in a prediction’s absolute error do not equate to changes in outcomes predicated on that forecast. Even when one forecast is more accurate than another, if both are beyond the utility horizon then the ”better” one is not actually more useful, just arbitrarily closer to the truth. This idea is the basis for the plateaued CRE point scoring function, which in turn is the basis for the WCIS. While a metric that does not always increase the penalty as forecast accuracy diminishes may seem counterintuitive, we believe that for characterizing contextual utility, a score with a limited scope of relevance is actually more intuitive than a score that gets arbitrarily worse (or better) no matter how far away it is from being helpful.

The WCIS builds on the Weighted Interval Score, adding the *δ*-parameterization to impel users to directly characterize contextual utility. Judging predictions in this way allows for a powerful and effective normalization of the error, making the WCIS easy to interpret and compare across heterogeneous forecasting scenarios. Importantly, this robust efficacy exists *only for each individual definition of utility*. We belabor this point because it is inherent to our overall assertion about forecast interpretability: that a specific use case is necessary to meaningfully evaluate prediction quality. Without an explicit link to how forecasts are used, there is no way to consistently and meaningfully evaluate them over variable spatial and temporal conditions. Other evaluation metrics are in essence arbitrary until they are contextualized, whereas the WCIS builds this contextualization directly into the formulation of the score.

## 5 Conclusions

Determining the future role of pandemic forecasting, as well as identifying areas of forecasting that need improvement, must at some point include the translation of modeling results to policy and decision makers. The WCIS is expressly intended to function well in this process, allowing for intuitive characterization of forecast utility that can be easily communicated to an audience with less technical expertise. Figure 3 demonstrates this directly. Without effective contextual normalization, generating such a display would be challenging given large differences in error magnitude, likely requiring a transformation (such as log-scaling) that limits interpretability. Instead, the WCIS allows for a direct, clearly defined interpretation of forecast utility to be displayed and compared in a technically meaningful and intuitively understandable way.

We created the WCIS to enable and encourage honest and contextually specific discourse about the utility of short-term epidemic predictions. It incorporates prediction uncertainty, keeps the technical definition of utility as simple as possible, and generates an intuitively interpretable and comparable numerical output. Our intent is to allow for people without specific technical experience to be able to interact with and evaluate probabilistic forecasting in a meaningful way. As the public health community learns from COVID-19 and prepares for future challenges, explicit analysis of the utility of historical predictions is essential. We hope the WCIS will help with effective and meaningful communication between modelers and practitioners in this effort.

## Data Availability

All data produced in the present study are available online at https://github.com/cpt-diabetes/wcis

https://doi.org/10.5281/zenodo.6301718

https://healthdata.gov/d/j4ip-wfsv

## Declarations

### Ethics approval and consent to participate

Not applicable. Ethics approval was not sought or required for this study. All human health data used are population-level COVID-19 outcomes sourced from publicly available online repositories.

### Consent for publication

Not applicable.

### Availability of data and materials

Code and processed data for both the facility- and state-level analyses are accessible from a publicly available GitHub repository [https://github.com/cpt-diabetes/wcis]. Forecast and ground truth data used for our state-level analysis are available from the COVID-19 Forecast Hub repository [https://doi.org/10.5281/zenodo.6301718]. The original source for ground truth hospitalization data is the COVID-19 Reported Patient Impact and Hospital Capacity by Facility repository [https://healthdata.gov/d/j4ip-wfsv].

### Authors’ contributions

MM conceived of the study, performed the analysis, and wrote the manuscript. MM and FP developed the methodology. LMG supervised the project and provided essential methodological guidance. All authors read, edited, and approved the final manuscript.

## Acknowledgements

Not applicable.

## Additional Files

### “Additional file 1.pdf”

This document contains the Supplemental Materials for this article. These include sections that provide more detail on and/or motivating examples for the formulation of the score, the impropriety analysis, and the facility-level model formulation. It also includes figures comparing the WIS and the WCIS performance of the facility-level model for different scenarios, and state-level WCIS hospitalization heatmaps for all 4 standard Forecast Hub prediction horizons (1, 2, 3, and 4 weeks ahead).

