## Additional file 1 for "When are predictions useful? A new method for evaluating epidemic forecasts"

### 1 Formulation

#### 1.1 Motivating Examples - Spatial and Temporal Variability

From many perspectives, making and disseminating state-level forecasts is a reasonable strategy. States are the intuitive building blocks of the country, carrying their own governments and public health systems. Accurate state-level forecasts therefore have the potential for direct and meaningful application. However, states have enormously variable characteristics, which makes generalizing forecast performance problematic. Population difference in particular is a key factor. For example, California has the highest population of any state in the US (∼40 million), and Wyoming the lowest (∼0.6 million). For the second week of January 2022, California reported over 850,000 incident cases. During the same week, Wyoming reported just over 6,600 new cases [1]. Note that California reported over 1.4 times more new cases that week than the entire population of Wyoming. However, in terms of incidence percentages, California and Wyoming were actually much closer at that time, with approximately 2% and 1% of the population testing positive, respectively. Intuitively, this is an easy dynamic to recognize when examining individual states separately. Raw epidemic numbers carry different meanings depending on underlying demographic factors (i.e., population size). However, this is problematic for aggregate and comparative analysis of forecast performance. This becomes clear if we apply a standard metric like mean absolute error (MAE) to this scenario with California and Wyoming. (For simplicity we refer to point predictions instead of probabilistic forecasts in the motivating examples in this section, along with corresponding metrics such as the absolute and percent error.

However, as indicated above, probabilistic evaluation is susceptible to the same issues [2].) For the week under consideration, predictions from the Forecast Hub's baseline model yielded a MAE of 27,130 across all US states [3]. For California, a prediction that overshot the truth by this margin would incur a percent error of only about 3%, whereas for Wyoming, such a prediction would miss by over 400%. Unfortunately, spatial inconsistency is not the only obstacle. Accounting for temporal context is equally vital and presents its own difficulties.

When examining forecast performance for a single region over time, metrics must be interpreted as a function of time-variant data. This necessity is demonstrated trivially by comparing pandemic surges to times of relatively low epidemic activity. The same value of a non-normalized metric like the absolute error carries an entirely different meaning in each of these situations. Consider the Forecast Hub's baseline model predictions for cases in Maryland. In mid-December 2020, this model missed its three-weeks-ahead target by about 2,000 cases. In mid-May 2021, the same model also missed by about 2,000 cases [3]. Without knowing the context of each prediction, (namely that the first was made during a massive surge and the second was made during a significant lull), one might be forgiven for assuming that the model performed similarly in both scenarios. However, the December forecast only just missed the mark, undershooting by 12% of the true value. Conversely, the May forecast missed by 213%. Note that in this case, percent error has interpretable utility because it normalizes by the true value, a time-varying data source that directly represents the prevailing condition of the pandemic. Unfortunately, percent error is not an ideal solution as it becomes unstable when true values approach zero [2]. This is especially problematic when analyzing death forecasts (for all of 2020 through 2022, almost 15% of US states had less than ten weekly deaths, and over 8% had below five weekly deaths). In this situation, percent error is in fact too sensitive to the exact circumstances. It indicates a relatively large deviation from the truth which, while technically correct, misses the reality of how forecasts are interpreted. Given the larger context of the pandemic, it is unreasonable to characterize a four-death forecast compared to a target value of one (300% error) as a worse prediction than a 400-death forecast compared to an 800-death reality (50% error). Like the spatial case, the numerical value of an error metric, absent any temporal contextualization, cannot be relied on to consistently or intuitively reflect forecast performance.

#### 1.2 Derivation of the CIS as a function of the CRE

For simplicity and consistency with extant metrics, we introduce the Contextual Interval Score (CIS) in the main body of the paper as a scaled, constrained version of the Interval Score (IS). However, we developed the CIS as a direct, interval-forecast extension of our point-forecast CRE function. In this section, we demonstrate that the formulation of the CIS as a function of the IS is equivalent to a different formulation that directly incorporates the CRE. Then, we explain each of the components of the equivalent form of the score to help intuit the motivation for the creation of the score.

We begin with the form of the CIS introduced in the main body of the paper:

$$CIS_\alpha(F, y, \delta) = min\left\{\frac{\alpha}{2\delta} IS_\alpha(F, y),\ 1\right\} \tag{1}$$

Taking the right hand side of this equation, we substitute in the expanded form of the Interval Score (IS):

$$min\left\{\frac{\alpha}{2\delta}\left[(u - l) + \frac{2}{\alpha}(l - y)\mathbb{1}\{y < l\} + \frac{2}{\alpha}(y - u)\mathbb{1}\{y > u\}\right],\ 1\right\} \tag{2}$$

Simplifying:

$$min\left\{\frac{\alpha}{2\delta}(u - l) + \frac{l - y}{\delta}\mathbb{1}\{y < l\} + \frac{y - u}{\delta}\mathbb{1}\{y > u\},\ 1\right\} \tag{3}$$

Examining the $\frac{l-y}{\delta}\mathbb{1}\{y < l\}$ term, we observe that if this term reaches or exceeds 1, the minimizer operating over the entire equation will restrict the overall output to 1. Thus applying a“local” minimizer, constraining this term to a maximum of 1, will not change the overall value of the score. The same logic applies to the the $\frac{y-u}{\delta}\mathbb{1}\{y > u\}$ term. Including these internal minimizers yields the following form of the CIS:

$$min\left\{\frac{\alpha}{2\delta}(u - l) + min\left\{\frac{l - y}{\delta},\ 1\right\}\mathbb{1}\{y < l\} + min\left\{\frac{y - u}{\delta},\ 1\right\}\mathbb{1}\{y > u\},\ 1\right\} \tag{4}$$

We can further exploit the indicator functions to include absolute values in the two minimized terms:

$$min\left\{\frac{\alpha}{2\delta}(u - l) + min\left\{\frac{|l - y|}{\delta},\ 1\right\}\mathbb{1}\{y < l\} + min\left\{\frac{|u - y|}{\delta},\ 1\right\}\mathbb{1}\{y > u\},\ 1\right\} \tag{5}$$

This equation now directly includes the formulation of the CRE. Thus, we can substitute the CRE in to show the complete alternate formulation of the CIS:

$$CIS_\alpha(F, y, \delta) = min \begin{Bmatrix} \frac{\alpha}{2\delta}(u-l) + CRE(l, y, \delta)\mathbb{1}\{y < l\} + CRE(u, y, \delta)\mathbb{1}\{y > u\} \\ 1 \end{Bmatrix} \tag{6}$$

Each term in the CIS is analogous to a term in the IS. We begin with the "width" term: $\frac{\alpha}{2\delta}(u-l)$. Because $y-\delta$ to $y+\delta$ represents the upper and lower limits of forecast utility, a prediction interval that spans this entire distance should incur an unweighted penalty of 1. In other words, if a point forecast at or past the "plateau" of the CRE curve incurs a penalty of 1, an unweighted interval forecast that spans this region should get the same score. However, the $\alpha$-weight is included to distinguish between different prediction intervals. Consider two intervals that have identical bounds but different $\alpha$ values: 0.05 (95% prediction interval) and 0.9 (10% prediction interval). In this case, the 95% interval should be treated less harshly that the 10% interval, because we expect higher-confidence forecasts to span larger ranges. Next, we examine the "miss" term of the CIS: $(CRE(l, y, \delta))\mathbb{1}(y < l) + (CRE(u, y, \delta))\mathbb{1}(y > u)$. It is essentially performing the same function as the "miss" term of the IS, but instead of expressing the magnitude of the miss in terms of distance, the CIS term is expressed in terms of utility. This component of the score can be seen in panels *(c)* and *(d)* of Additional file 1: Fig. S1 as the vertical arrows. In sum, the CIS is a single-interval analogue of the point-forecast CRE. Regardless of interval width, if a probabilistic forecast is entirely outside the useful region, a value of 1 is returned (panel *(d)* in Fig. S1). Like the IS, the CIS naturally collapses to only its "miss" term when applied to a point forecast.

#### 1.3 Visualization of the CRE and CIS

The following figure, S1, provides a visualization of the CRE and the three different ways the CIS can arise, depending on the relative positions of the prediction interval bounds and the true value. Panel *(a)* shows only the Contextual Relative Error (CRE) point score (Equation 3 in the main text), with the others displaying different realizations of the Contextual Interval Score (CIS, Equation 4 in the main text). Blue arrows represent the width penalty term (note that they are scaled by $\frac{\alpha}{2\delta}$). Red arrows indicate the miss term of the CIS. Observe that because the miss term is not scaled, any forecast that entirely misses the $y-\delta$ to $y+\delta$ region, regardless of width, will incur the maximum penalty of 1. For clarity, each of the panels refers to a single-interval evaluation. The full Weighed Contextual Interval Score (WCIS) is composed of an average across multiple $\alpha$ intervals.

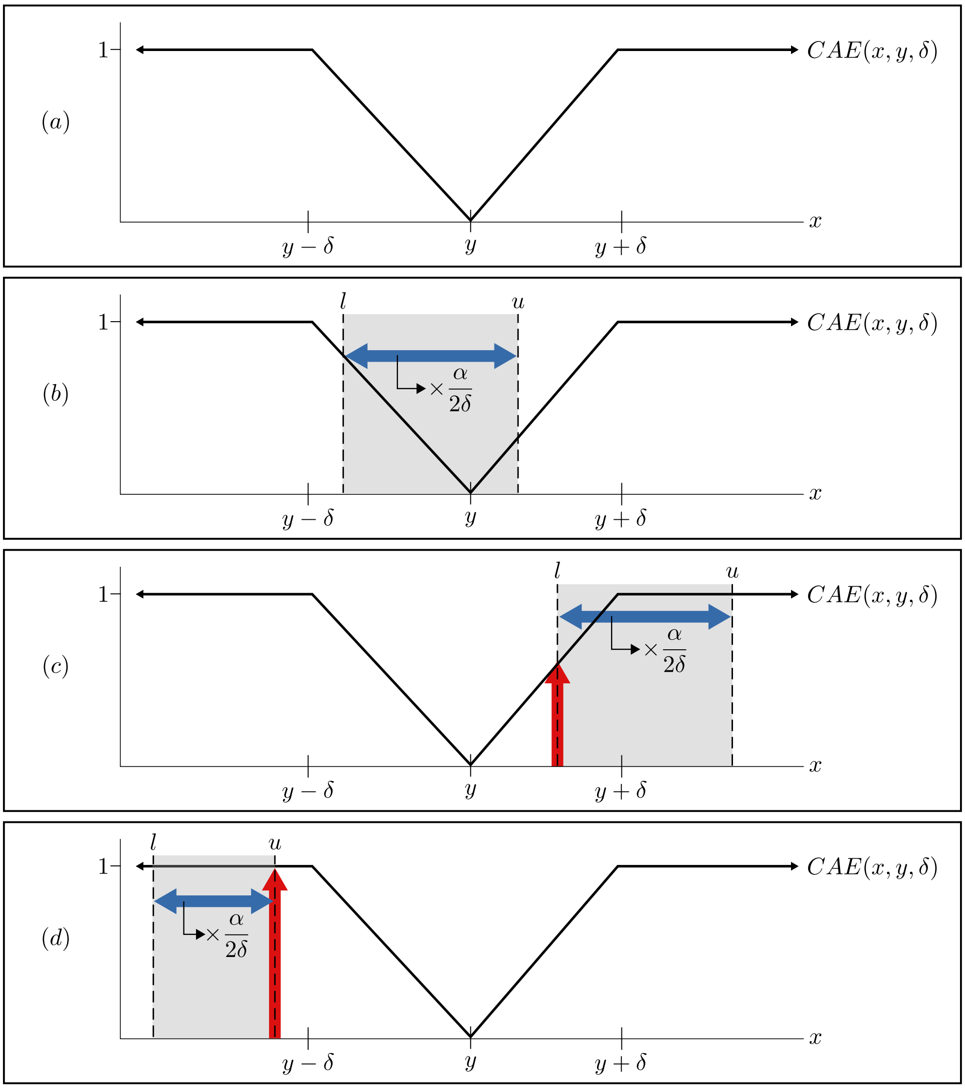


$$CIS_{\alpha}(F, y, \delta) = min\left\{ \tfrac{\alpha}{2\delta} \boxed{(u-l)} + \boxed{CAE\,(l, y, \delta)\,\mathbb{1}\,\{y < l\} + CAE(u, y, \delta)\mathbb{1}\,\{y > u\}},\ 1 \right\}$$

**Fig. S1** Demonstration of the CRE (Panel a) and the three different calculation modes that the CIS can take (Panels b,c,d).

#### 1.4 Empirical Impropriety Demonstration

In this test case, we select an arbitrary distribution to represent the output of a forecasting model. This distribution functions as the source of the modeler's "good-faith" predictions, i.e. a proper score will incentivize submission of forecasts that are legitimately representative of this distribution. Next, we select an arbitrary $\delta$-parameterization and interval represented by $\alpha$. "Good-faith" predictions are drawn as the $\frac{\alpha}{2}$ and $1 - \frac{\alpha}{2}$ quantiles of the distribution. Next, we iterate over the domain of the distribution and take the expected score for each feasible interval pair. If any pair has a score that is lower in expectation than the "good-faith" interval, then the score is not proper.

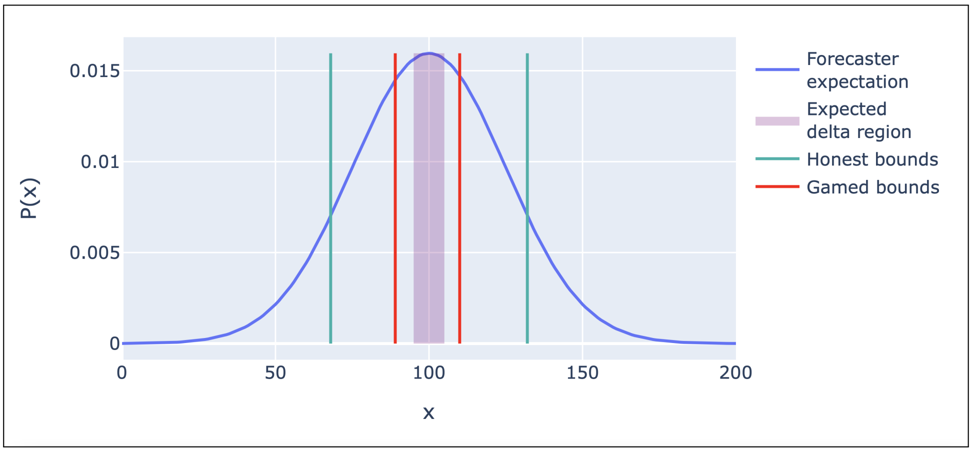


**Fig. S2** Empirical demonstration of the impropriety of the Contextual Interval Score (CIS). Note the difference between the "honest" and "gamed" interval bounds, indicating an incentive to deviate from submitting good-faith statistical realizations in a effort to minimize the expected penalty. The parameterization here is a normal distribution defined with $\mu = 100$ and $\sigma = 25$. $\delta$ and $\alpha$ were selected to be 5 and 0.2, respectively.

As is clearly demonstrated by figures S2 and S3, the CIS and therefore the multi-interval WCIS is not a statistically proper interval score. However, we propose that a score with the desired features of the WCIS is inherently improper. The foundation of the WCIS is the notion of a specific and *constrained* region around the target value wherein predictions are applicable, represented by the V-shaped CRE function. This means that from a gaming/error minimization perspective, the WCIS could encourage probabilistic forecasts that are affected by the size of the $\delta$-region [4]. Similar to prior forecasting efforts when improper metrics were used, propriety is sacrificed in exchange for other, desirable properties of the score [5–7]. Additionally, ongoing work by Bosse et al. indicates that applying monotonic transformations like the natural logarithm to target data can help to alleviate the domination of higher-activity forecasting scenarios for model comparison and aggregation while retaining propriety [8].

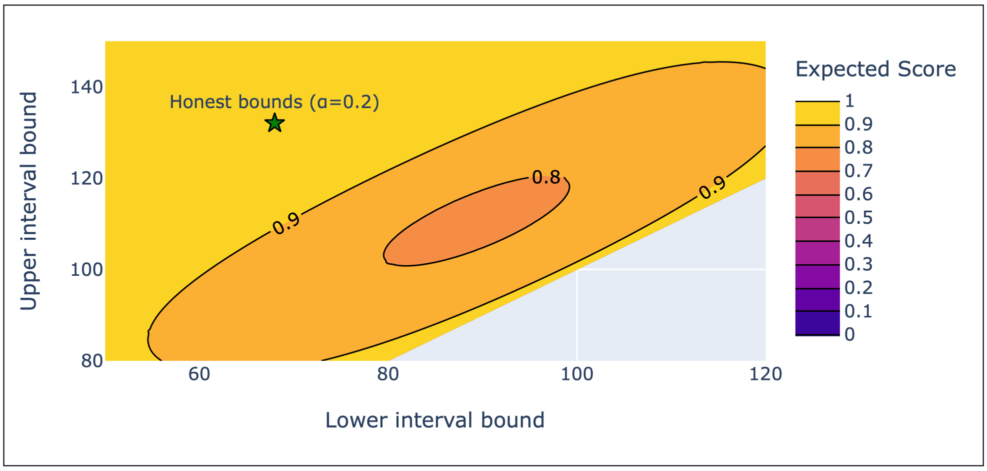


**Fig. S3** Contour plot showing the expected Contextual Interval Score (CIS) value for a set of different prediction interval bounds. The distribution and score parameterization used here is the same as in Figure S2 above. Note that the "honest" bounds do not exist at the minimum expected score, and selecting bounds that do minimize the expectation results in the "gamed" bounds (approximately 89 and 110) that are seen in Figure S2.

### 2 Facility-Level Analysis

#### 2.1 Facility-Level Model Formulation

**Data:** We obtained facility-level data on COVID-19 hospitalizations from the COVID-19 Reported Patient Impact and Hospital Capacity by Facility dataset, collected at various times by the CDC, HHS, and CDC again. This dataset includes metrics related to COVID-19 hospitalizations, hospital occupancy, and capacity [9]. For this analysis, we focused only on the time series of COVID-19 bed occupancy. The raw data was provided at a weekly resolution for each hospital. To enable more granular modeling, we performed temporal disaggregation to obtain daily resolution data. We assumed that the weekly trends at each hospital followed the same pattern as the aggregated state-level trends, which were available at a daily resolution. For each week, we normalized the daily state-level values to sum to 1, then multiplied the normalized values by each hospital's weekly totals to impute daily hospital-level values. Any remaining missing values were imputed using local regression smoothing [10]. We selected 42 hospitals in Maryland for this analysis. Hospitals were included if they were classified as short-term acute care hospitals and if they had at least 10 COVID-19 patients at some point between July 2021 and July 2022. We chose to focus on a single state because modeling and analyzing all U.S. hospitals was not practical.

**Model:** To forecast future COVID-19 hospitalizations, we used the Time Series Dense Encoder (TiDE) model, a deep neural network architecture that has achieved state-of-the-art performance on general time-series forecasting tasks [11]. TiDE uses a simple but flexible encoder-decoder structure that can incorporate covariates and accommodate various prediction horizons, output distributions, and loss functions.

We configured the TiDE model with 4 encoder layers, 4 decoder layers, a decoder output dimension of 32, hidden size of 128, past temporal width of 4, future temporal width of 4, dropout probability of 0.1, and layer normalization. The model used the previous 90 days as context to predict hospitalizations for the next 21 days. Rather than making sequential autoregressive predictions, the model predicted all 21 days at once. To obtain probabilistic forecasts, we used quantile regression, with the model directly outputting predictions for the 0.01, 0.025, 0.05, 0.1, 0.15, 0.2, 0.25, 0.3, 0.35, 0.4, 0.45, 0.5, 0.55, 0.6, 0.65, 0.7, 0.75, 0.8, 0.85, 0.9, 0.95, 0.975, and 0.99 quantiles. The input features were the target variable (total COVID-19 census) and time covariates including year, day of year, day of week, and days since July 1, 2021. Predictions were generated for each Monday in the time span included, simulating a weekly system like the Forecast Hub's [3].

**Training and Calibration:** We trained a separate model for each hospital and prediction date using an expanding window of training data starting from August 1, 2020. Models were trained for 100 epochs to minimize the pinball loss. The model predictions were post-processed in two steps to improve calibration. First, we applied a non-negativity constraint, thresholding all predictions to be at least 0 since negative patient counts are impossible. Second, we applied the conformalized quantile regression (CQR) method [12]. CQR adjusts the predicted quantiles based on the model's historical quantile errors to achieve better coverage. We did not hold out a separate calibration dataset, instead using the training data for the CQR calibration.

**Implementation:** We implemented the models in Python using the darts time-series library, while the data processing was done in Julia [13]. Model training took approximately 10 seconds per hospital and prediction window using an NVIDIA 4070Ti GPU. ß

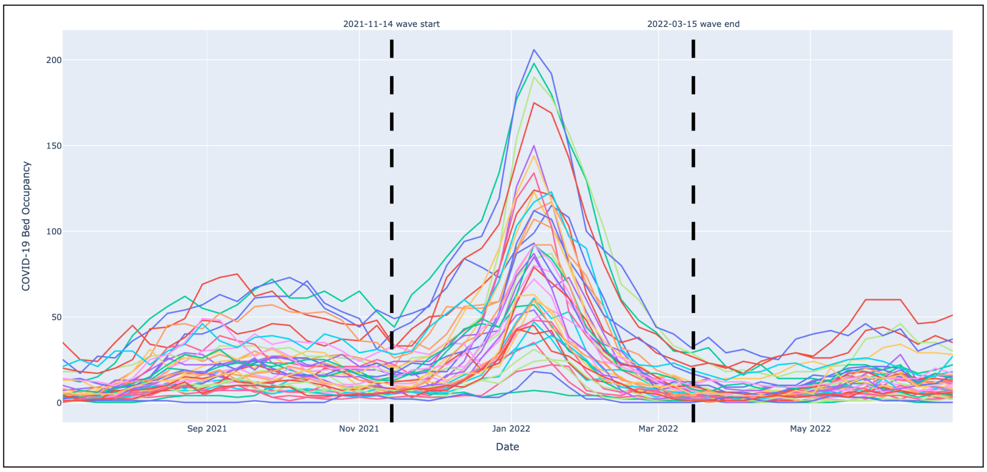


**Fig. S4** Delineation of the segments of the facility-level analysis that are in and out of the Omicron wave for the purposes of our analysis. Each line represents one of the 42 Maryland facilities predicted for.

**Table S1** Hospitals Included in Facility-Level Model

| Hospital ID | Facility Name |
|---|---|
| 210001 | Meritus Medical Center |
| 210002 | University of Maryland Medical Center |
| 210003 | University of Maryland Prince George's Hospital Center |
| 210004 | Holy Cross Hospital |
| 210005 | Frederick Health Hospital |
| 210006 | University of Maryland Harford Memorial Hospital |
| 210008 | Mercy Medical Center |
| 210009 | The Johns Hopkins Hospital |
| 210011 | Saint Agnes Hospital |
| 210012 | Sinai Hospital of Baltimore |
| 210015 | MedStar Franklin Square Medical Center |
| 210016 | Adventist Healthcare White Oak Medical Center |
| 210017 | Garrett County Memorial Hospital |
| 210018 | MedStar Montgomery Medical Center |
| 210019 | TidalHealth Peninsula Regional, Inc. |
| 210022 | Suburban Hospital |
| 210023 | Anne Arundel Medical Center |
| 210024 | MedStar Union Memorial Hospital |
| 210027 | U.P.M.C. Western Maryland |
| 210028 | MedStar Saint Mary's Hospital |
| 210029 | Johns Hopkins Bayview Medical Center |
| 210032 | Union Hospital of Cecil County |
| 210033 | Carroll Hospital Center |
| 210034 | MedStar Harbor Hospital |
| 210035 | University of Maryland Charles Regional Medical Center |
| 210037 | University of Maryland Shore Medical Center at Easton |
| 210038 | University of Maryland Medical Center Midtown Campus |
| 210039 | CalvertHealth Medical Center |
| 210040 | Northwest Hospital Center |
| 210043 | University of Maryland Baltimore Washington Medical Center |
| 210044 | Greater Baltimore Medical Center |
| 210048 | Howard County General Hospital |
| 210049 | University of Maryland Upper Chesapeake Medical Center |
| 210051 | Doctors Community Hospital |
| 210056 | MedStar Good Samaritan Hospital |
| 210057 | Adventist Healthcare Shady Grove Medical Center |
| 210060 | Adventist Healthcare Fort Washington Medical Center |
| 210061 | Atlantic General Hospital |
| 210062 | MedStar Southern Maryland Hospital Center |
| 210063 | University of Maryland St. Joseph Medical Center |
| 210064 | Levindale Hebrew Geriatric Center And Hospital |
| 210065 | Holy Cross Germantown Hospital |

#### 189 2.2 Facility-Level Metric Comparisons

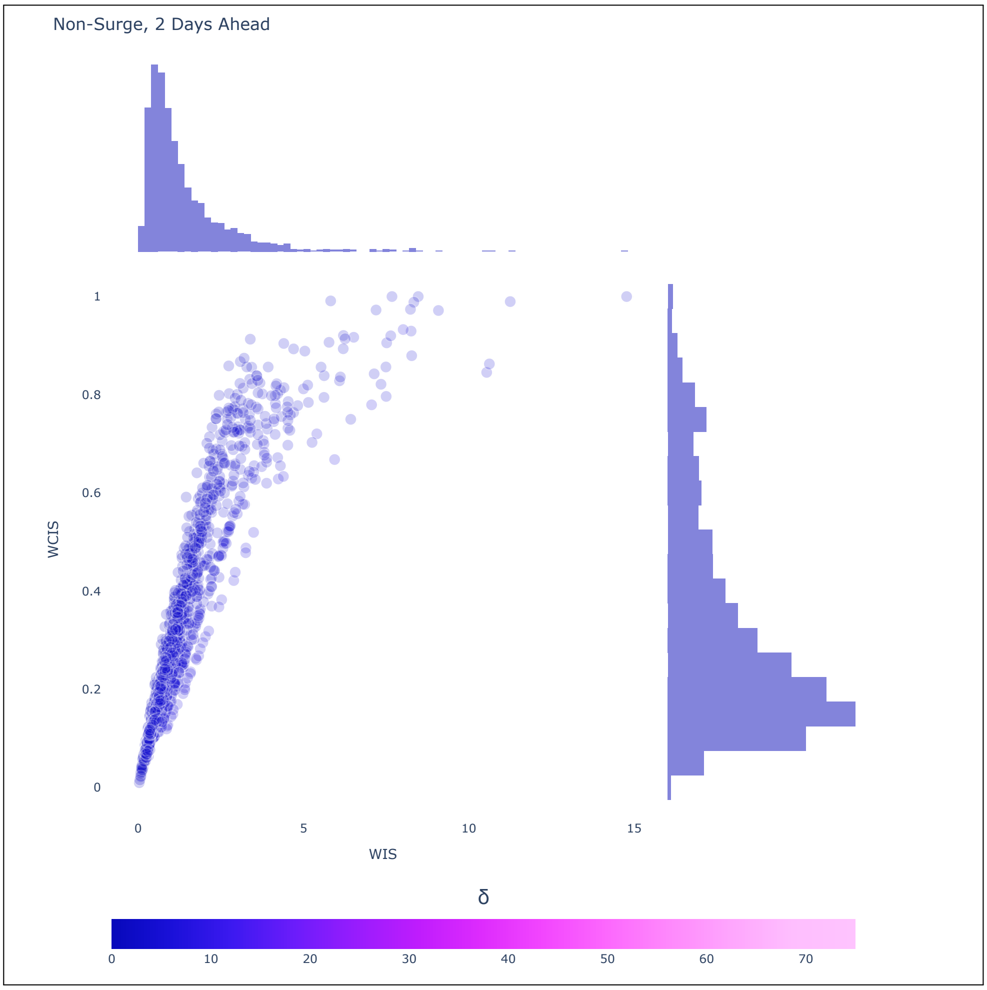


**Fig. S5** WIS vs WCIS values for all 42 facilities, for 2-day-ahead forecasts, for all prediction dates outside of the Omicron surge.

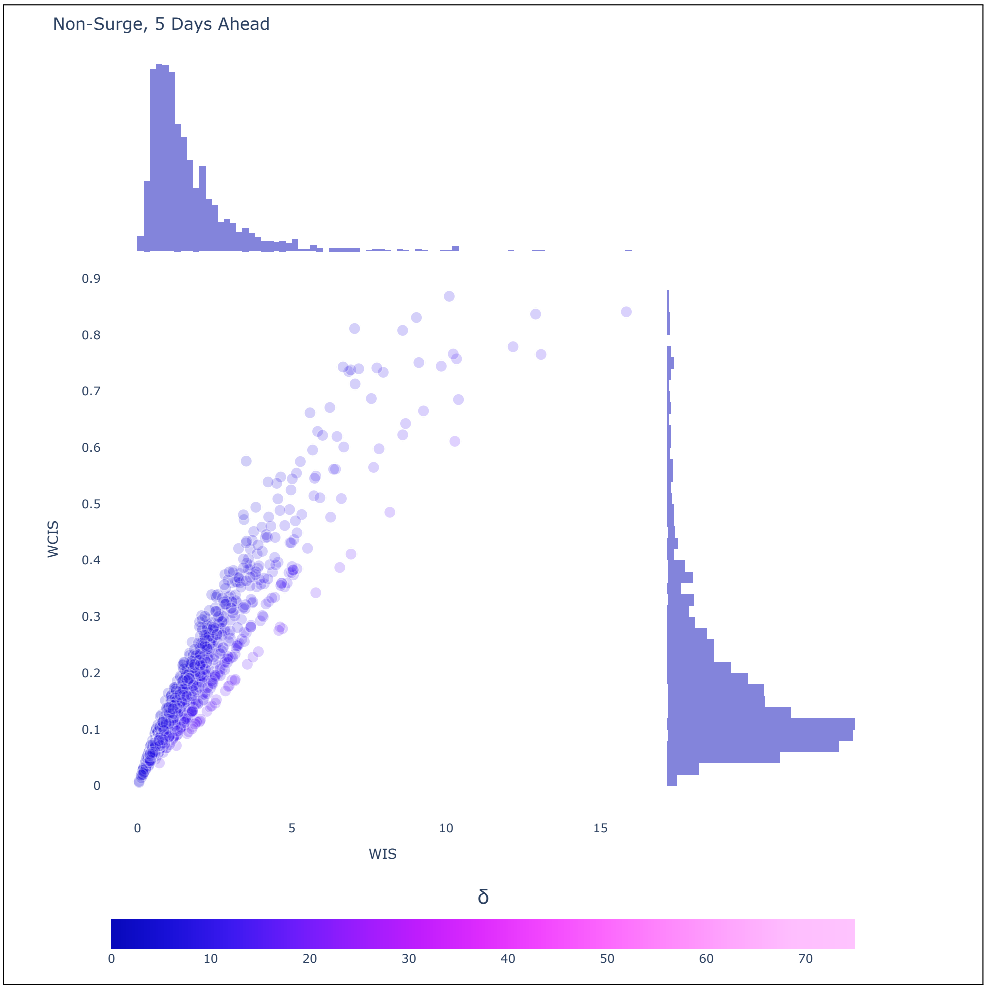


**Fig. S6** WIS vs WCIS values for all 42 facilities, for 5-day-ahead forecasts, for all prediction dates outside of the Omicron surge.

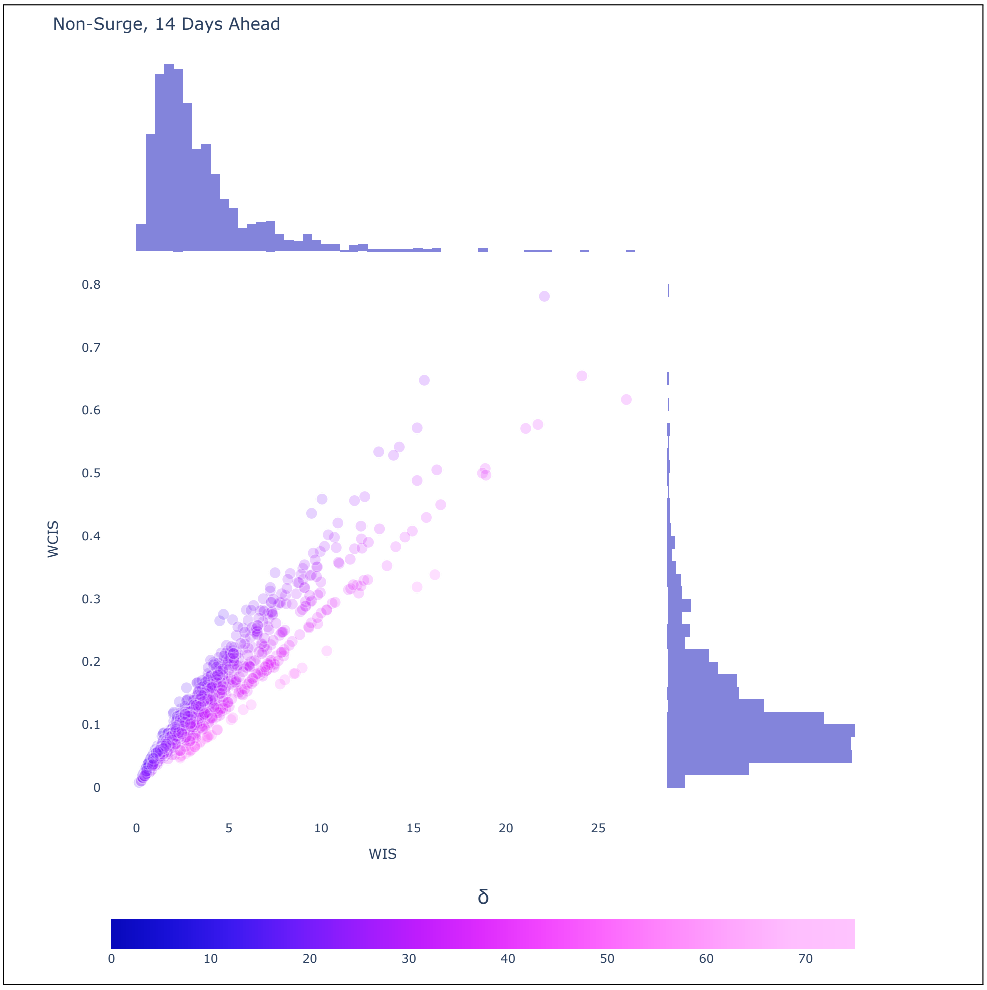


**Fig. S7** WIS vs WCIS values for all 42 facilities, for 14-day-ahead forecasts, for all prediction dates outside of the Omicron surge.

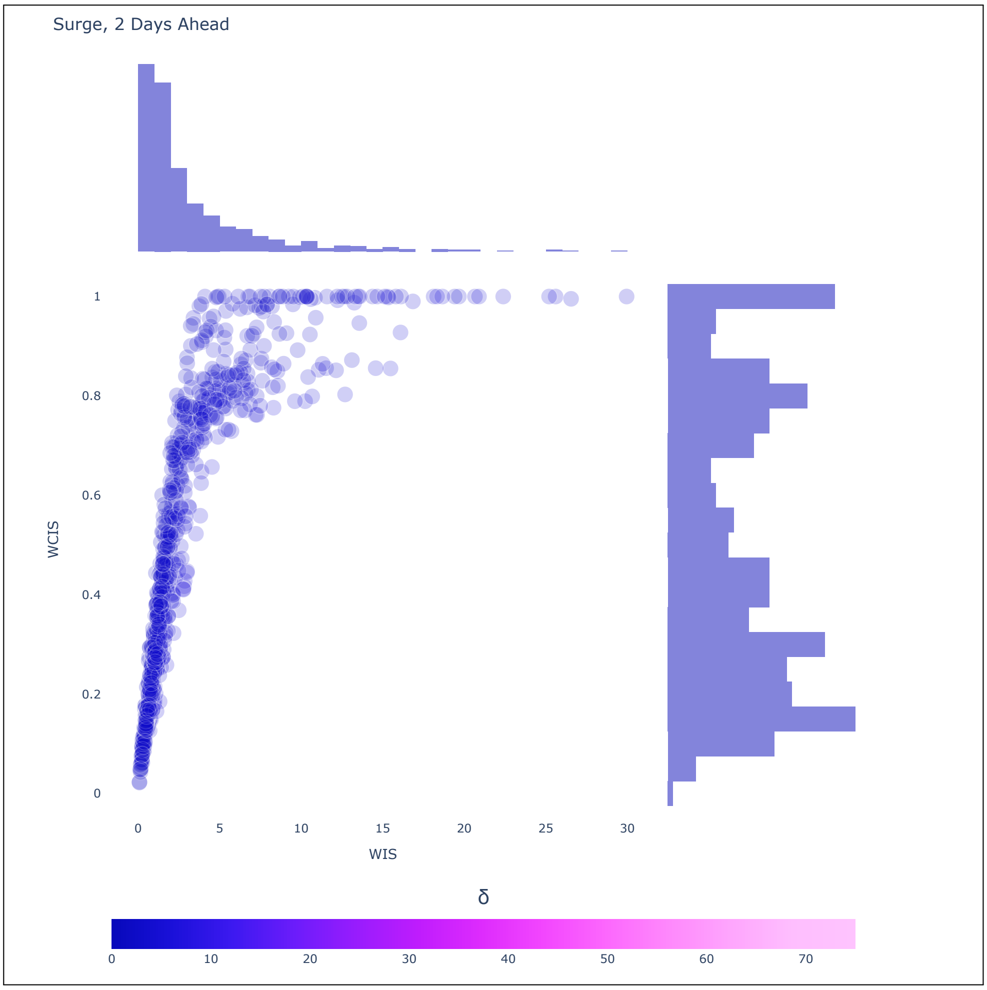


**Fig. S8** WIS vs WCIS values for all 42 facilities, for 2-day-ahead forecasts, for all prediction dates within the Omicron surge.

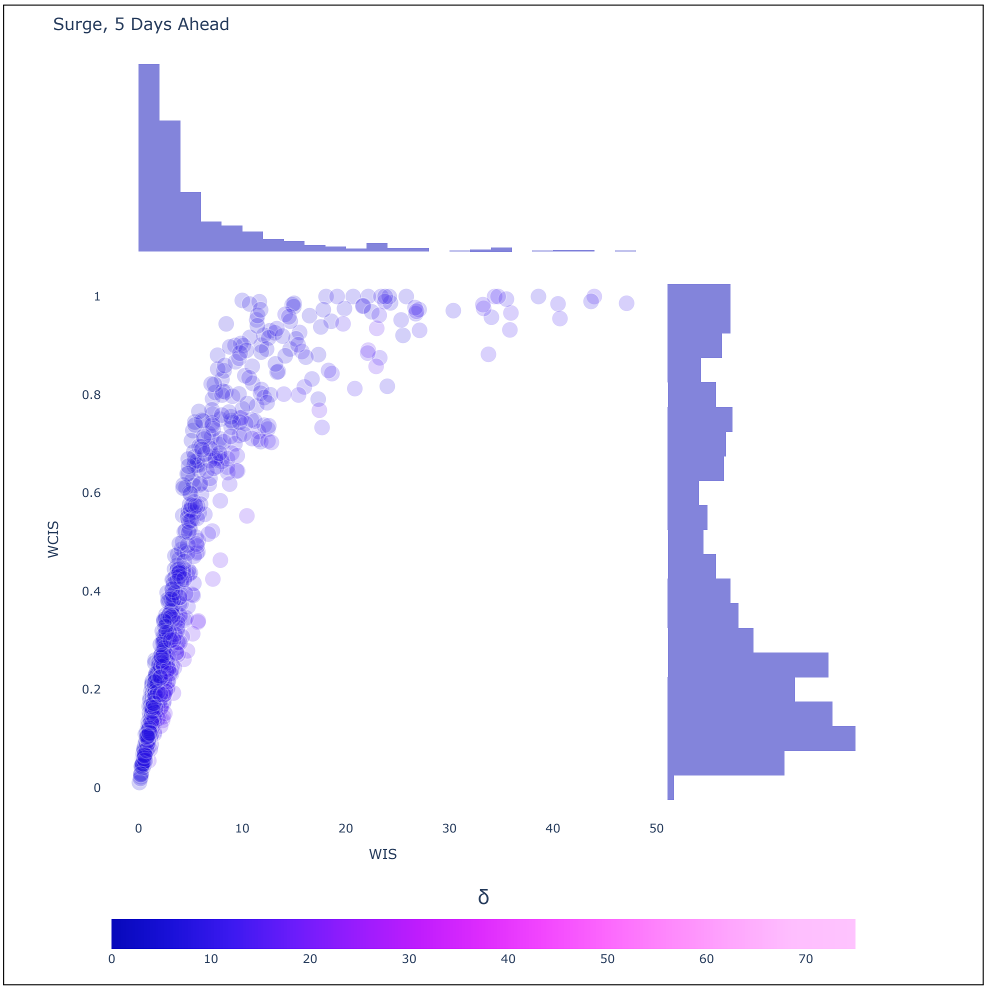


**Fig. S9** WIS vs WCIS values for all 42 facilities, for 5-day-ahead forecasts, for all prediction dates within the Omicron surge.

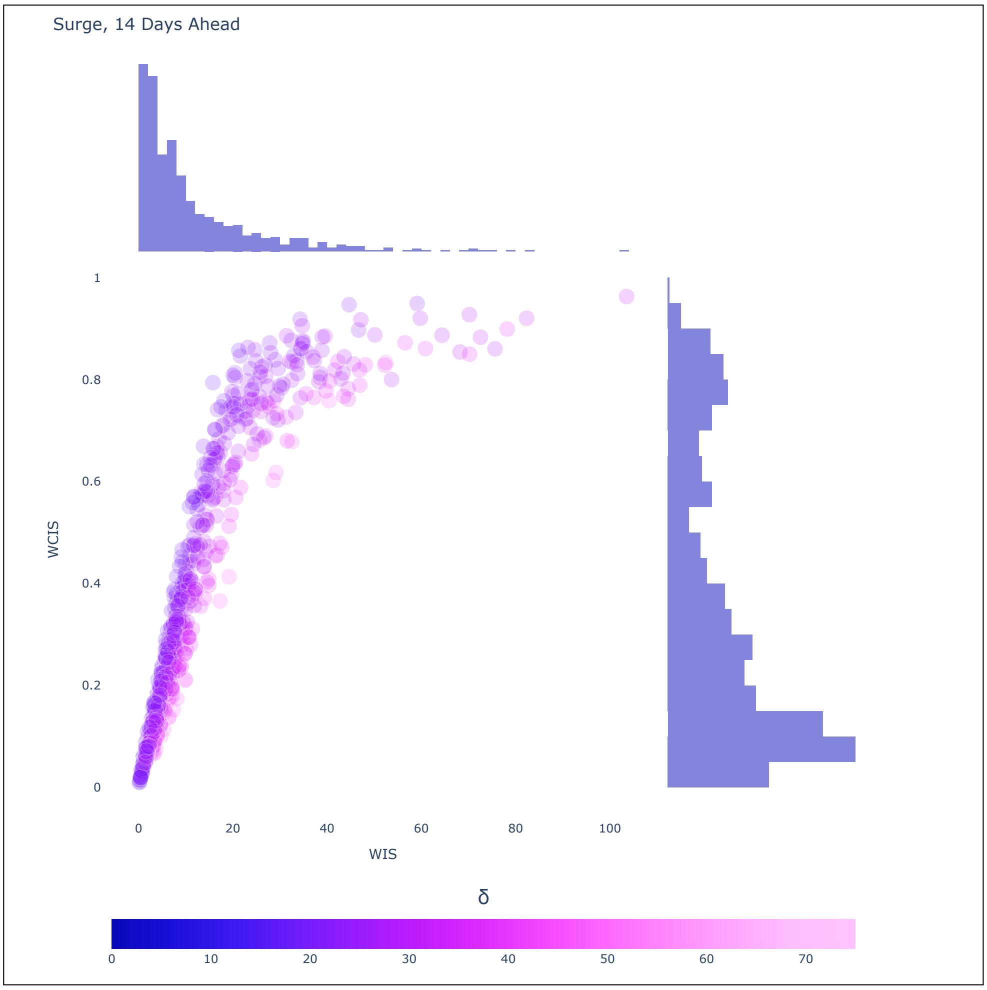


**Fig. S10** WIS vs WCIS values for all 42 facilities, for 14-day-ahead forecasts, for all prediction dates within the Omicron surge.

### 3 COVIDhub Ensemble Hospitalization Forecasts (Second Test Case)

Included here are heatmaps of the WCIS vs the WIS for hospitalization forecasts for each prediction horizon (one, two, three, and four weeks ahead) from the Forecast Hub's ensemble model. The $\delta$ used for the hospitalization analysis is detailed in full in section 3.2 of the main text of the paper. We note here that the column used to generate the delta values is "inpatient_beds" in the COVID-19 Reported Patient Impact and Hospital Capacity by Facility dataset (archive link: https://healthdata.gov/d/j4ip-wfsv).

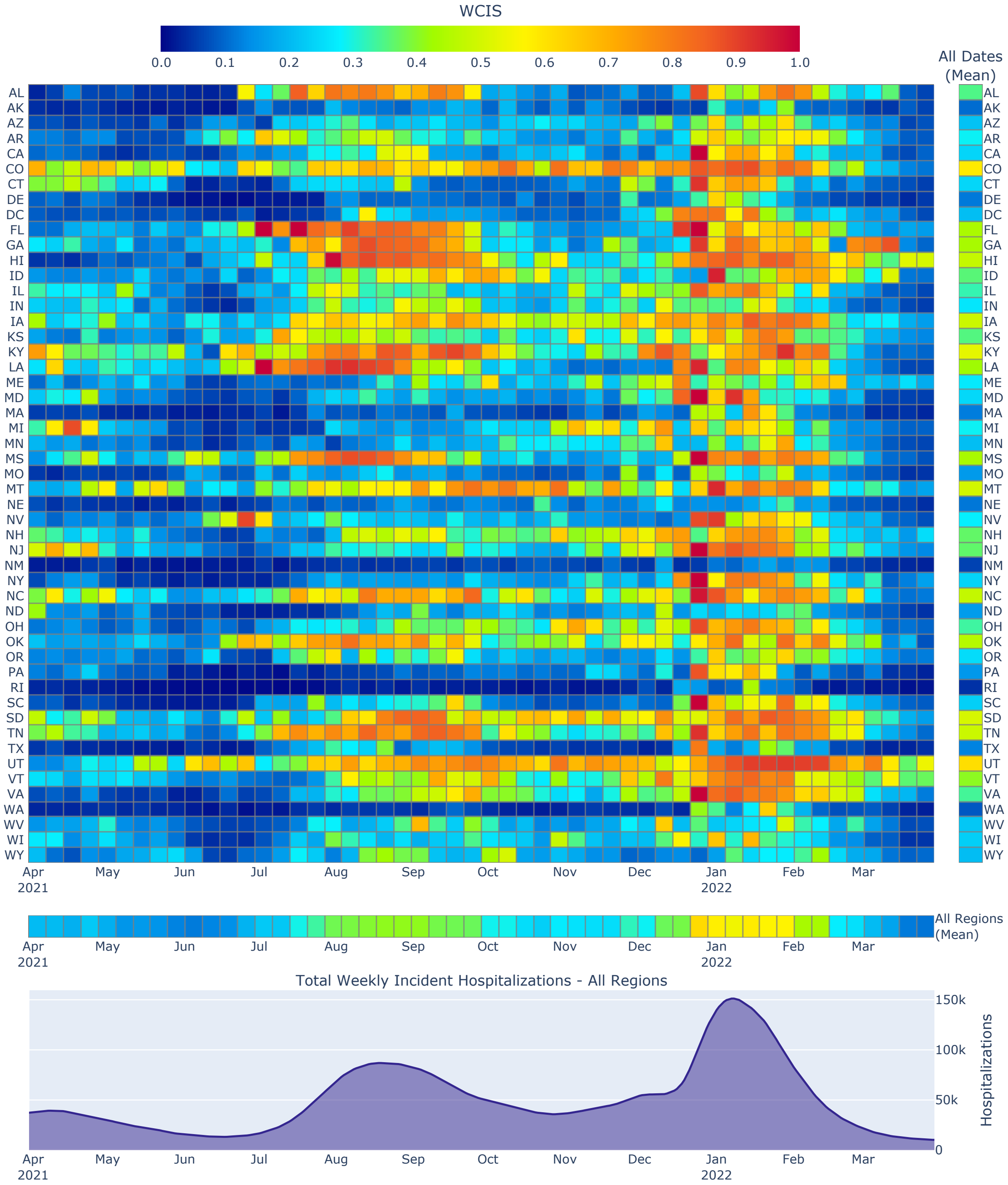


**Fig. S11** Heatmap of the WCIS for 1 week ahead hospitalization forecasts, performed by the Forecast Hub's ensemble model.

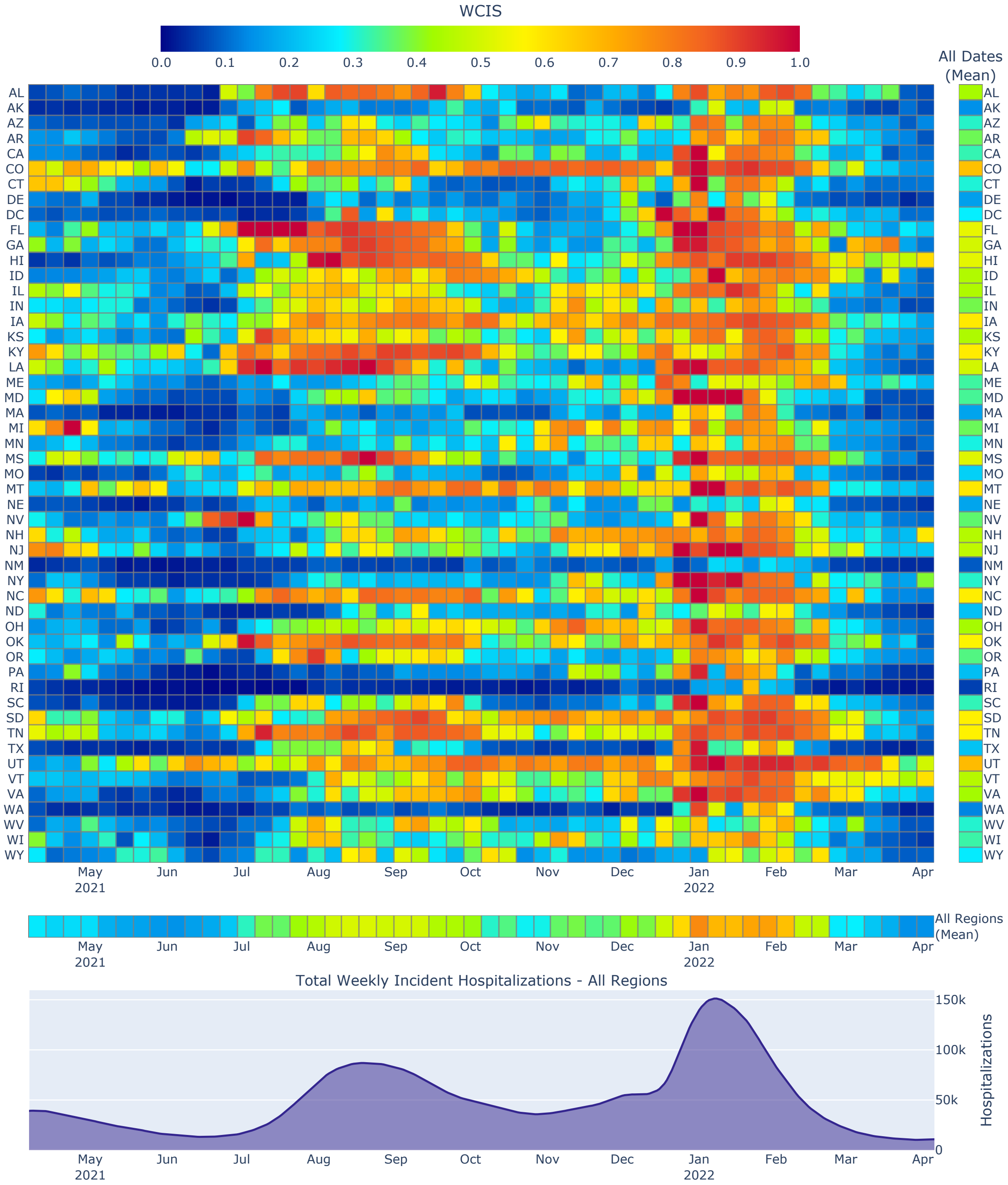


**Fig. S12** Heatmap of the WCIS for 2 week ahead hospitalization forecasts, performed by the Forecast Hub's ensemble model.

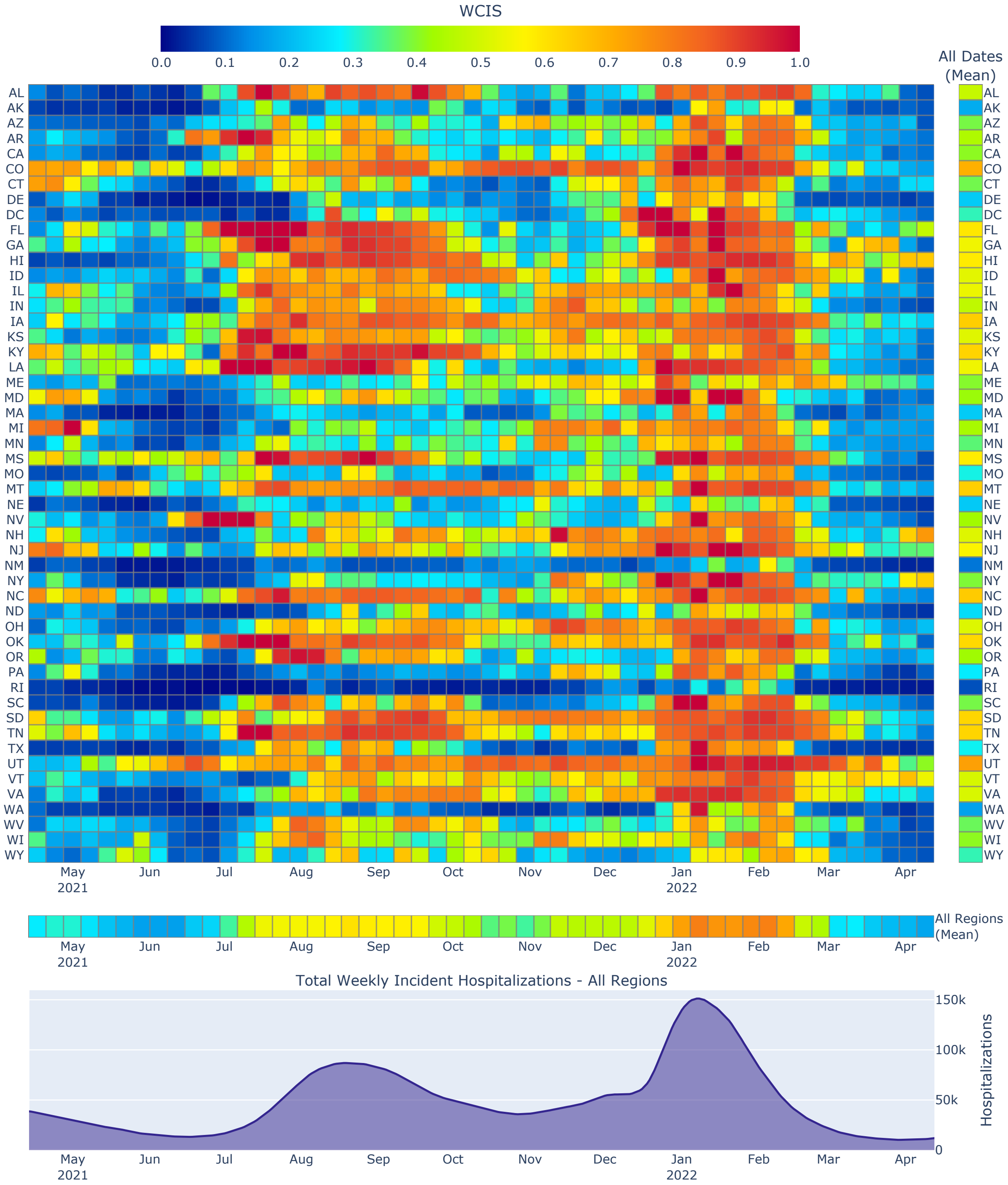


**Fig. S13** Heatmap of the WCIS for 3 week ahead hospitalization forecasts, performed by the Forecast Hub's ensemble model.

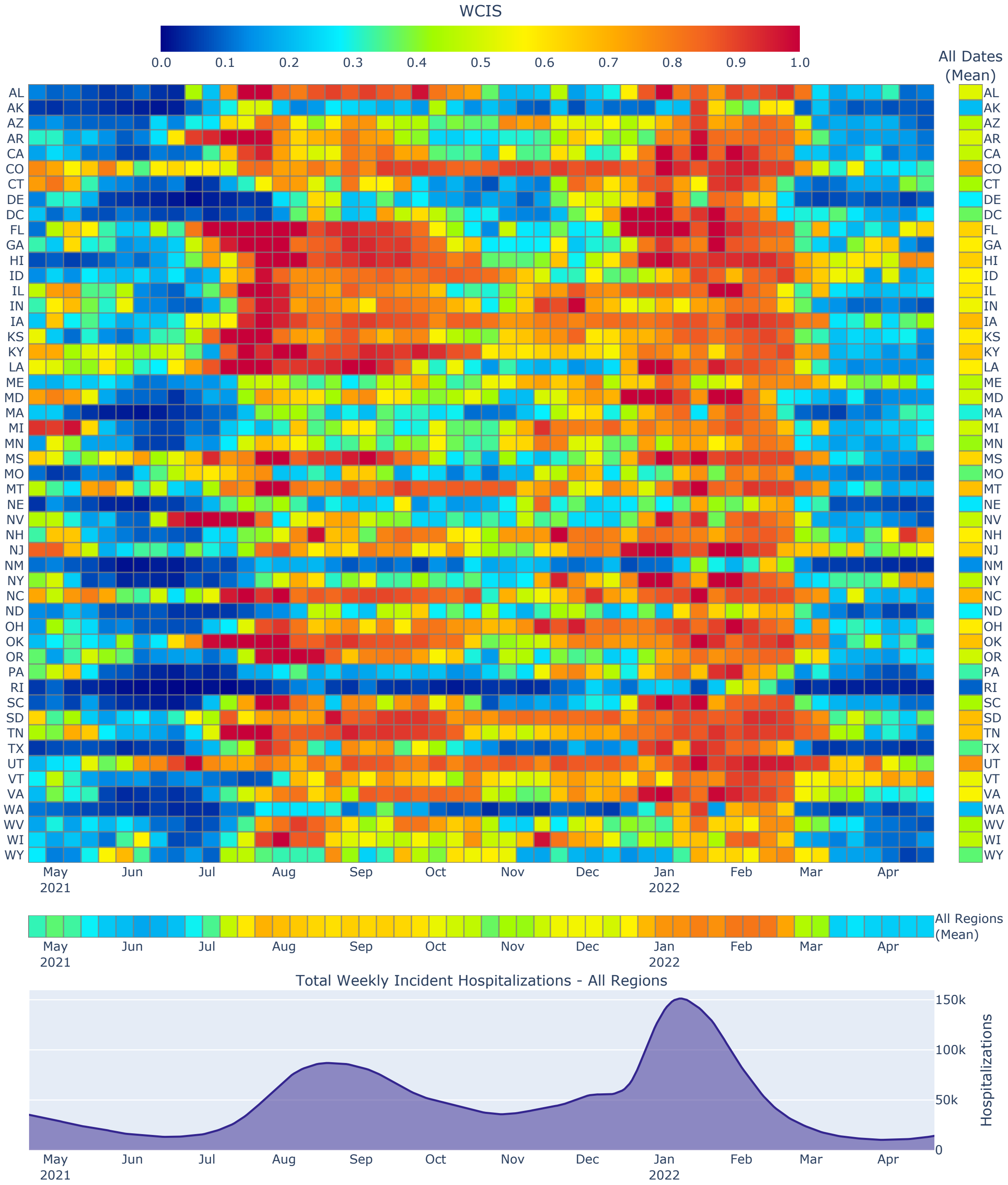


**Fig. S14** Heatmap of the WCIS for 4 week ahead hospitalization forecasts, performed by the Forecast Hub's ensemble model.
